# A slow road from meat dominance to more sustainable diets: an analysis of purchase preferences

**DOI:** 10.1101/2021.11.25.21266850

**Authors:** Maijaliisa Erkkola, Satu M Kinnunen, Henna R Vepsäläinen, Jelena M Meinilä, Liisa Uusitalo, Hanna Konttinen, Hannu Saarijärvi, Mikael Fogelholm, Jaakko Nevalainen

**Affiliations:** Department of Food and Nutrition, University of Helsinki, Helsinki, Finland; Faculty of Social Sciences, University of Helsinki, Helsinki, Finland; Faculty of Management and Business, Tampere University, Tampere, Finland; Health Sciences, Faculty of Social Sciences, Tampere University, Tampere, Finland

**Keywords:** grocery purchases, protein source, transition, sociodemographic, price

## Abstract

**Background:** Achieving a sustainable and healthy diet requires increased replacement of red meat with more sustainable foods. There is a call for novel methodologies to assess the potential of different interventions and policies in enhancing the transition from the current to more sustainable choices.

**Objective:** We aimed to characterize consumer clusters with similar preferences in protein sources, to compare the purchase prices of these foods, and to identify ongoing transitions from one protein source to another.

**Design:** Grocery purchase data with individual attributes on 29,437 consenting loyalty card holders were analyzed over 2.3 year period. We designed a sequence analysis to group participants to clusters with similar purchase preferences over the follow-up period and to estimate transition probabilities between preferences. We studied the determinants of prevalent purchase profiles by ordinal logistic models.

**Results:** We identified six participant profiles with similar preferences in four protein sources: red meat, poultry, fish, and plant-based foods. Red meat dominated the purchase preferences and showed the highest persistence over time. The majority (70%) of the participants demonstrated somewhat mixed purchase profiles. A step-by-step transition from red meat towards plant-based food preference seems most likely via poultry and fish. Overall, low income was not a barrier to a more sustainable purchase profile, while price may deter the purchase of fish. The most important resources in choosing more sustainable profiles were education and stage of family life.

**Conclusions:** Societal incentives for sustainable food choices seem most crucial at transition stages of life course and for the less educated. Here we also demonstrate that grocery purchase data offer a valuable tool for monitoring the progressive transition towards a healthy and sustainable food system.

## Introduction

In the past century, a massive increase has occurred in the consumption of animal-based products, with a nearly doubled quantity of meat available for worldwide consumption between 1961 and 2017 (1, 2). This global carnivore approach is neither nutritionally nor environmentally sustainable (3, 4). There is an increasing endorsement of a new global standard, a more plant-based diet, to achieve the 2030 United Nations sustainable goals and to relieve the burden of non-communicable diseases (5-9). One of the key claims of the Planetary Health Diet was that by 2050 the global consumption of red meat will need to reduce by more than 50% from the 2019 level (8). Achieving a sustainable diet would entail increased replacement of red meat with sustainable fish and plant-based alternatives and a simultaneous increase in fruit and vegetable intake (3, 6, 10, 11). However, both extremes – strict animal product avoidance or high intake of red and processed meat – pose nutritional risks (3, 11-15).

Multiple and complex issues influence consumers’ attitudes and behavior towards meat consumption. Consumers should be able to distinguish between accurate information and misinformation shaping their knowledge and attitudes (16). While making choices, consumers may consider taste preferences, personal and family health, diet-related environmental issues, animal welfare, religious or other ideologies, and cost (17-21). The weight of these issues varies by consumers’ characteristics. Earlier studies show that meat eaters are more likely to be men, middle-aged, less educated, and with a family (17,18, 22, 23).

There is a call for multiple indicators and novel methodologies to assess the potential of different interventions, policies, and social media debates in enhancing the transition from the current to more sustainable dietary choices (24, 25). We used the extensive individualized purchase data of the leading grocery retailing loyalty card program in Finland, with the *first aim* of identifying and characterizing consumer clusters with similar preferences in protein sources. While red meat remains the most important meal component in most high-income countries, transitions away from red meat as the main dish towards poultry, fish, and plant-based foods were examined. Our *second aim* was to compare the purchase prices of these foods. To use the detailed data efficiently, we undertook a customized sequence analysis of the preferred protein sources over time, enabling our *third aim*: the identification of ongoing transitions from one protein source to another. We emphasized the role of everyday, educational, and economic resources and demographic factors in sustainable purchase behavior and considered nutrition literacy and attitude as possible mediators. Given that dietary behaviors are modifiable and most people consume these protein sources on a daily basis, our findings have critical public health and environmental implications.

## Methods

### Study design

Data were obtained from the S Group, which is the largest grocery retailer in Finland, with a market share of 46% in 2018 (26). Members of the S Group’s loyalty card program are provided with a customer card to be used when making purchases, and loyalty card holders are rewarded for their purchases by receiving a maximum 5% monetary reward that is monthly refunded to them. Households’ primary card holders across Finland were contacted via email and invited to participate in the study, which involved consenting to the release of their grocery purchase data for research purposes and voluntarily responding to an online questionnaire that included questions on resources, attitudes, nutrition literacy, special diets, and degree of loyalty to a retailer chain. The study was approved by the University of Helsinki Ethical Review Board in the humanities and social and behavioral sciences (Statement 21/2018). Each participant provided an informed consent electronically.

### Participants

Initially, 47,066 loyalty card holders consented to release their 2.3-year grocery purchase data (Supplemental Figure 1). We did not have information on the number of valid email addresses or the proportion of emails reaching the card holders (e.g. bypassing trash email filters). Of the participants, 36,621 (78%) responded to the online questionnaire. For inclusion, we set a threshold of self-estimated degree of loyalty at ≥41%; the threshold value was based on an earlier observation that the relative shares of purchases of main food groups were similar among these participants (27). Thus, 29,437 participants were included in the analyses.

### Background characteristics

Card holders’ sex and age were obtained from the retailer’s database. Information on resources (family structure, education, household income) was collected via the questionnaire. The participants were classified into five family structure categories: single-adult households, one adult and a child/children, two adults, two adults and a child/children, or other (households with three or more adults and households with unknown family structure). Participants reported their education on a four-point scale: primary school or less, upper secondary school, lower level tertiary, or higher level tertiary. In addition, monthly gross household income was reported on a seven-point scale ranging from less than 1,500 € to 9,000 € or more. Scaled monthly household income was then calculated as the mean income in each of the categories divided by the square root of household size (OECD square root scale) and classified into five categories: less than 1,000 €/month, 1,000–1,999 €/month, 2,000–2,999 €/month, 3,000–3,999 €/month, and 4,000 €/month or more.

### Attitudes, nutrition literacy, and special diets

Attitudes and nutrition literacy were assessed with two questions measuring agreement with guidelines on fish, meat, and pulses given in the Nordic Nutrition Recommendations (28). Respondents were asked to rate their opinion on the statements “To promote health, the consumption of fish should be increased and that of red meat and processed meats limited” (attitude) and “Recommended dietary changes include increasing the consumption of pulses” (nutrition literacy). For both questions, a seven-point scale (from “1=fully disagree” to “7=fully agree”) was presented. We asked the respondents to indicate whether at least one member of their household followed a diet with no red meat or a vegetarian or vegan diet.

### Grocery purchase variables

The grocery purchase data used in this study covered the period from 1 September 2016 to 31 December 2018. Each purchase was linked to the card holder and associated with an item description, time stamp, and quantity (i.e. weight, volume, or number of packages) and cost of the item. The data consisted of 4,234 grocery product groups of which foods were re-grouped by a professional nutritionist. We used the following food groups: *red meat and processed meat (R)* including also processed white meat, *poultry and poultry dishes (P), fish and seafood (F)*, and *plant-based foods (PB)* including plant-protein products and vegetable dishes, but excluding vegetables as such. The purchases were expressed in kilograms per week.

### Statistical analysis

Grocery purchase data arise in a different way from traditional dietary data collection. Thus, we analyzed the data in a way that condenses information in an effective and interpretable way. Sequence analysis (29) is a technique for visualizing and learning from time-ordered categorical data. Here, we designed a sequence analysis to group participants to clusters with similar purchase preferences over the follow-up period and to estimated transition probabilities between preferences.

Differences in household size and in the degree of loyalty hinder comparisons between participants’ absolute purchase volumes (27). Therefore, we first conducted a within-participant comparison of products. For each week of the follow-up, we computed the purchased volume of each food group. As food purchased in one week was not necessarily consumed that week, purchase volumes were defined as a backward moving average of two consecutive weeks. We labeled the participants’ ‘purchase preference’ by the most purchased food group in that week, i.e. either as red meat, poultry, fish, or plant-based foods. Thus, each participants’ preference data could be expressed as a 123-letter long sequence, each letter representing the purchase preference at one week. Missing purchase preferences, due to absence of transactions in the two-week period, were coded as a preference of their own.

We defined the first week of September 2016 as the time origin and one week as the unit of time in the sequence analysis. To estimate *prevalent purchase preferences*, participants were clustered based on their temporal sequences of weekly preferences using the optimal matching criteria, which is a way to measure (dis-)similarities between sequences of categorical events, along with the hierarchical Ward clustering algorithm. Distances between preferences were defined based on observed transition rates in the data. The choice of the number of clusters was based on the height of consecutive steps in the dendrogram as well as on their interpretability; after six clusters, the qualitative meanings of the clusters started to overlap.

We investigated the determinants of prevalent purchase profiles by logistic (cumulative logit link) models with cluster membership as an ordinal outcome. To achieve a meaningful spectrum of sustainability of purchase profiles, clusters were ordered from the least sustainable to the most sustainable profile. “Sustainability” was defined by the proportion of purchased red meat in the profile. Thus, odds ratios >1 from this model indicated that the factor studied tended to associate with more sustainable purchase profiles. Due to the large sample size, we regarded statistical hypothesis tests as meaningless and focused on estimation of parameters characterizing the effect sizes. The proportional odds assumption was investigated by the Bayesian Information Criteria comparing models with equal and unequal parameters for each factor; no substantive deviations from the assumption were found.

Models were used to assess the role of demographic factors, resources, attitude, and nutrition literacy in the sustainability of purchase profiles. We fitted an unadjusted model that included factors one by one, a demographic and resource factors model, and a full model that also included attitude and nutrition literacy variables.

We estimated the purchase prices of food groups by purchase profile; we computed the sums of the expenditure (€) on and the volume (kg) of food groups for each participant. Means and standard deviations were estimated from the distribution of their quotients such that each participant had the same weight in the calculation. The same was done for the totals over food groups and profiles. Purchase prices per food product were derived by a direct division of the total expenditure by the total volume across participants, as all products were not purchased by all participants.

To detect *changes in purchase preferences*, we estimated the transition probabilities of switching from one preference (e.g. red meat and processed meat on week *k*) to another preference (e.g. fish and seafood on week *k+1*). We focused on persistence of preferences (i.e. no transition), transitions away from red meat and processed meat preference, and transitions to plant-based food preference.

Data were analysed using the TraMineR package in R (30) and SAS version 9.4.

## Results

Nearly one-third (29%) of participants representing their households lived with children, 56% had at least a lower level tertiary education, and 31% had a modal scaled household income of 2000-2999 €/month (Table 1). For 9%, household income was lower than 1000 €/month. Sixty-six percent of participants were women and 55% were at least 45 years old (Table 2).

**Table 1.** Overall distribution of resources and the distribution of purchase profiles for each level of resources (n=29 437).

|  | Overall | Red meat | Red meat slightly mixed | Red meat and poultry | Mixed | Fish | Plant-based | Cramer's V |
| --- | --- | --- | --- | --- | --- | --- | --- | --- |
|  | n (%) |  |  |  |  |  |  |  |
| Education |  |  |  |  |  |  |  |  |
| Primary school or less | 1878 (6%) | 747 (40%) | 771 (41%) | 69 (4%) | 250 (13%) | 16 (1%) | 25 (1%) | 0.125 (p<0.0001) |
| Upper secondary school | 10,994 (37%) | 3427 (31%) | 4685 (43%) | 829 (8%) | 1544 (14%) | 127 (1%) | 382 (3%) |  |
| Lower level tertiary | 9482 (32%) | 2143 (23%) | 4159 (44%) | 929 (10%) | 1583 (17%) | 179 (2%) | 489 (5%) |  |
| Higher level tertiary | 7010 (24%) | 1036 (15%) | 2834 (40%) | 678 (10%) | 1671 (24%) | 254 (4%) | 537 (8%) |  |
| Missing | 73 |  |  |  |  |  |  |  |
| Income €/month* |  |  |  |  |  |  |  |  |
| < 1000 | 2540 (9%) | 486 (19%) | 974 (38%) | 228 (9%) | 543 (21%) | 49 (2%) | 260 (10%) | 0.071 (p<0.0001) |
| 1000 - 1999 | 4373 (16%) | 1400 (32%) | 1835 (42%) | 319 (7%) | 590 (13%) | 41 (1%) | 188 (4%) |  |
| 2000 - 2999 | 8512 (31%) | 2276 (27%) | 3577 (42%) | 743 (9%) | 1356 (16%) | 160 (2%) | 400 (5%) |  |
| 3000 - 3999 | 6543 (24%) | 1664 (25%) | 2821 (43%) | 571 (9%) | 1072 (16%) | 121 (2%) | 294 (5%) |  |
| 4000 or more | 5477 (20%) | 1048 (19%) | 2455 (45%) | 473 (9%) | 1135 (21%) | 155 (3%) | 211 (4%) |  |
| Missing | 1992 |  |  |  |  |  |  |  |
| Family structure |  |  |  |  |  |  |  |  |
| 1 adult | 7441 (27%) | 1442 (19%) | 3020 (41%) | 685 (9%) | 1467 (20%) | 250 (3%) | 577 (8%) | 0.081 (p<0.0001) |
| 1 adult + children | 1300 (5%) | 397 (31%) | 573 (44%) | 128 (10%) | 125 (10%) | 14 (1%) | 63 (5%) |  |
| 2 adults | 10,343 (37%) | 2412 (23%) | 4391 (42%) | 834 (8%) | 2021 (20%) | 208 (2%) | 477 (5%) |  |
| 2 adults + children | 6546 (24%) | 1906 (29%) | 2969 (45%) | 539 (8%) | 865 (13%) | 62 (1%) | 205 (3%) |  |
| Other | 2196 (8%) | 723 (33%) | 884 (40%) | 187 (9%) | 321 (15%) | 24 (1%) | 57 (3%) |  |
| Missing | 1611 |  |  |  |  |  |  |  |

\* Scaled monthly household income calculated as the mean income in each of the categories divided by the square root of household size (OECD square root scale)

**Table 2.**
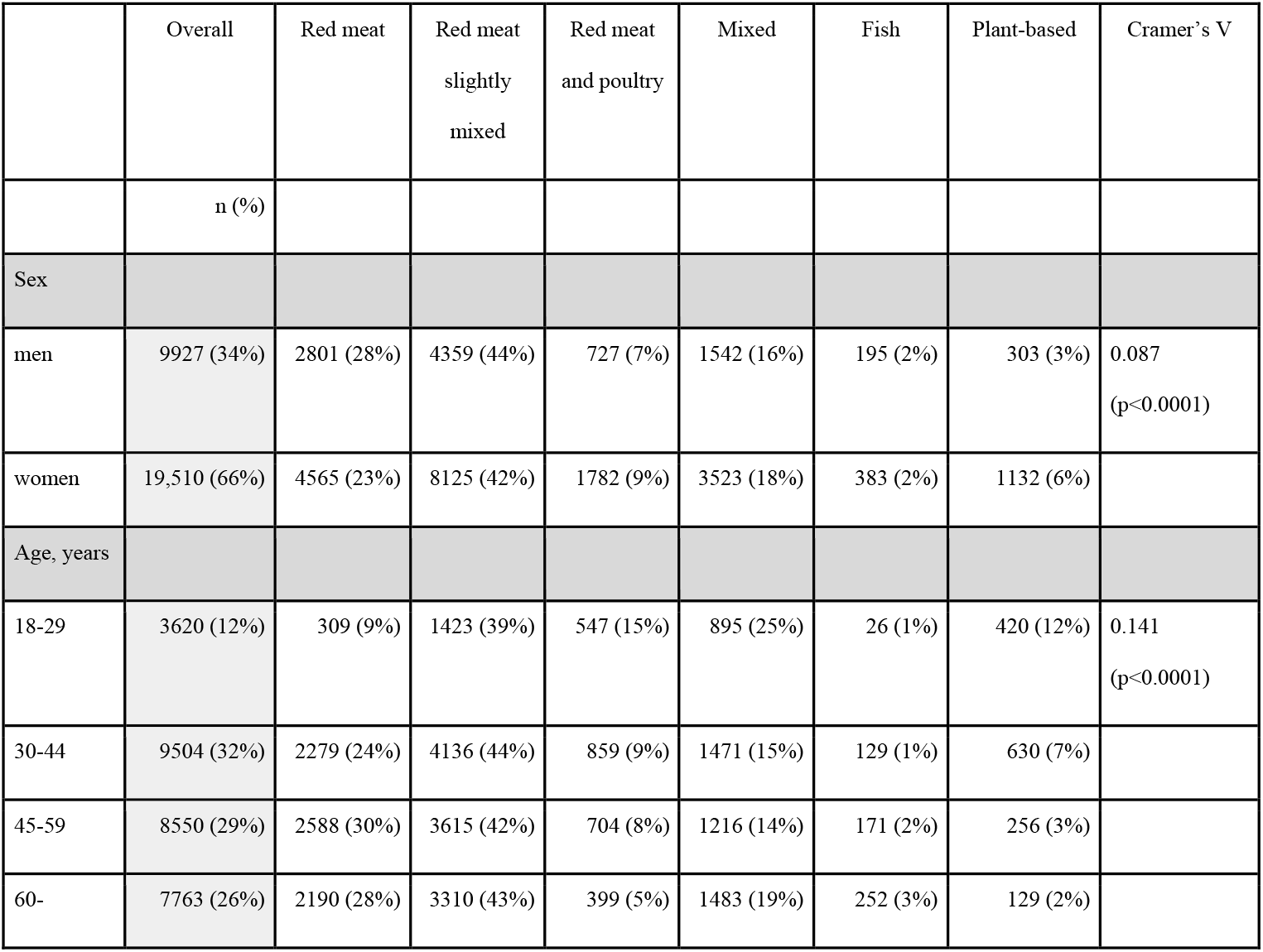
Overall distribution of demographic factors and the distribution of purchase profiles by demographic factors (n=29 437).

### Prevalent purchase preferences

Red meat and processed meat were by far the most common weekly purchase preference, accounting for 63% of main protein sources, followed by poultry and poultry dishes (14%), fish and seafood (11%), and plant-based foods (8%). Four percent of preferences were missing.

Six clusters, or purchase profiles, were identified by the sequence analysis. The largest cluster (“Red meat slightly mixed”) contained 42% of participants. Their primary purchase preference was red meat, but they also made regular purchases of poultry and fish, as seen from their combined share of close to one-third of the purchase volume. The second largest cluster (“Red meat”, 25%) displayed a more dominant preference towards red meat, with other alternatives together representing only one-quarter of the purchases. The third largest cluster, still of notable size (“Mixed”, 17%), displayed varying preferences. Proportions were more equally spread across food groups, ranging from 14% to 44%. The three smallest clusters had rather specific profiles: one (“Red meat and poultry”, 9%) purchased red meat and poultry in approximately equal amounts, but fish and plant-based products to a lesser extent. Participants with the “Plant-based” profile (5%) mostly preferred plant-based foods. Interestingly, poultry did not appear in the purchase profile nearly at all, and the proportion of fish was small. The smallest cluster consisted of 2% of the participants; most of their purchases were fish and seafood (“Fish”), followed by small but similar shares of red meat and plant-based foods. Profiles are shown in the order from least sustainable (highest proportion of red meat) to most sustainable (lowest proportion of red meat) (Figure 1).

**Figure 1.**
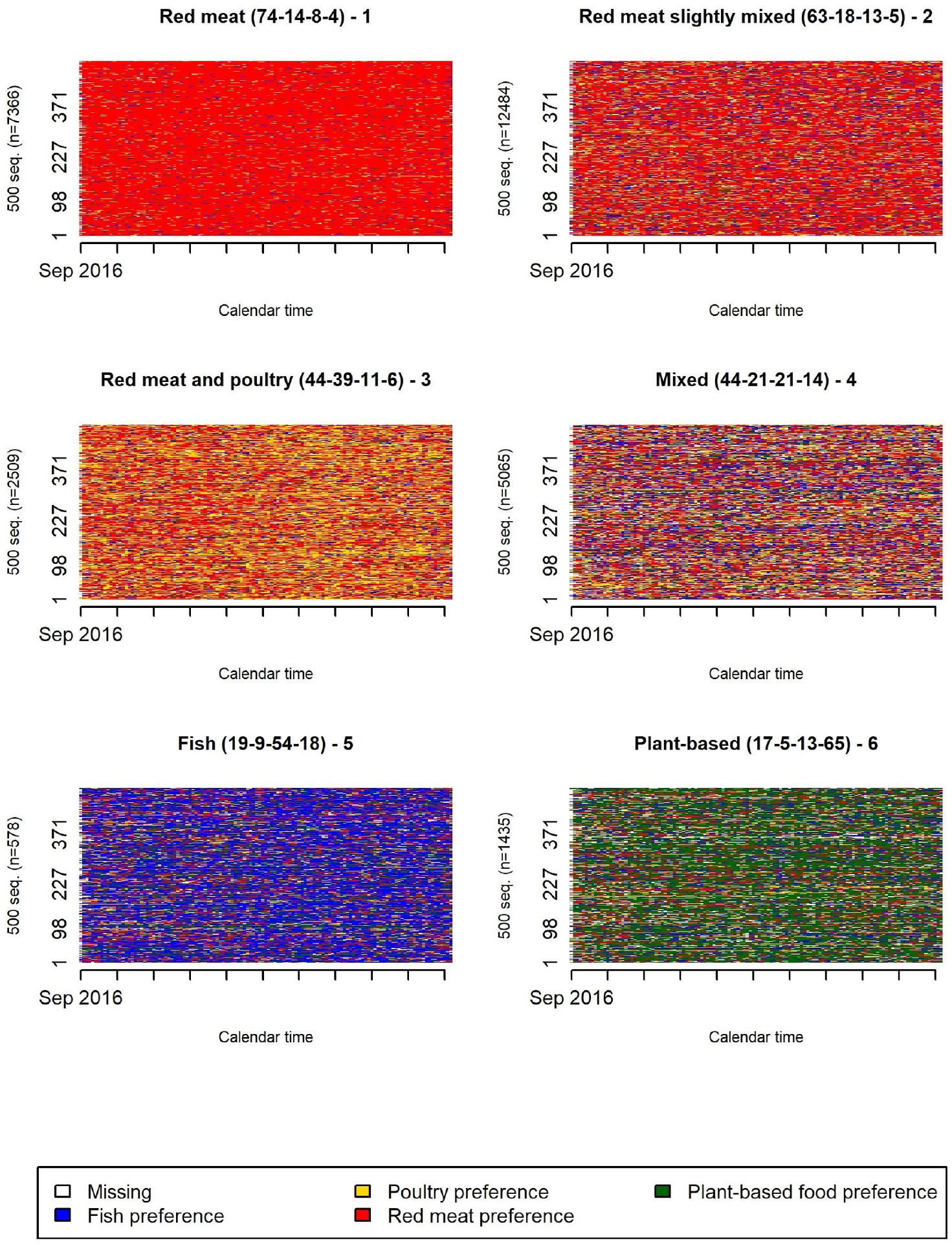
Purchase profiles identified by the sequence analysis, sorted from the least to the most sustainable. Each row in a panel represents an individual sequence of preferences with each week assigned the colour of the preference; 500 sequences per cluster are shown. The proportion of each food group (% of volume [kg] R-P-F-PB) is shown in the title. R = red meat and processed meat, P = poultry and poultry dishes, F = fish and seafood, and PB = plant-based foods.

### Purchase profiles and background characteristics

Table 1 shows the distribution of purchase profiles by participants’ resources. Among them, differences across education levels were the most pronounced; higher education was associated with a lesser likelihood of red meat dominance and with a greater prevalence of mixed, red meat and poultry, plant-based, and fish profiles. Of those with primary school (or less) education, 81% fell into clusters with red meat or red meat slightly mixed profiles. Participants from households with the lowest and the highest income levels were less likely to have a red meat dominant profile than those from other households. Plant-based profiles were the most frequent among those with the lowest household income. Households with children tended to prefer red meat more often than those without children.

A larger proportion of men had a red meat profile than women, and the opposite was observed for the plant-based profile (Table 2). The most striking difference between age groups was the profile distribution of the youngest age groups; only one in ten had the red meat dominant profile, and more than half had a mixed, red meat and poultry, or plant-based profile.

The frequency distributions of attitudes, nutrition literacy, and self-reported special diets by purchase profile are shown in Figure 2. Overall, 83% of participants agreed (response scoring ≥ 5) and 8% disagreed (response scoring ≤ 3) with the claim that the consumption of fish should be increased and that of red meat and processed meat decreased. The corresponding figures for an increase in consumption of pulses were 64% and 15%. Attitudes towards the recommended changes in diet tended to be positive in all clusters, but mostly so for clusters with mixed, plant-based food, or fish profiles. Participants with these profiles also reported the highest rates of vegetarian diets and red meat avoidance (Figure 2).

**Figure 2.**
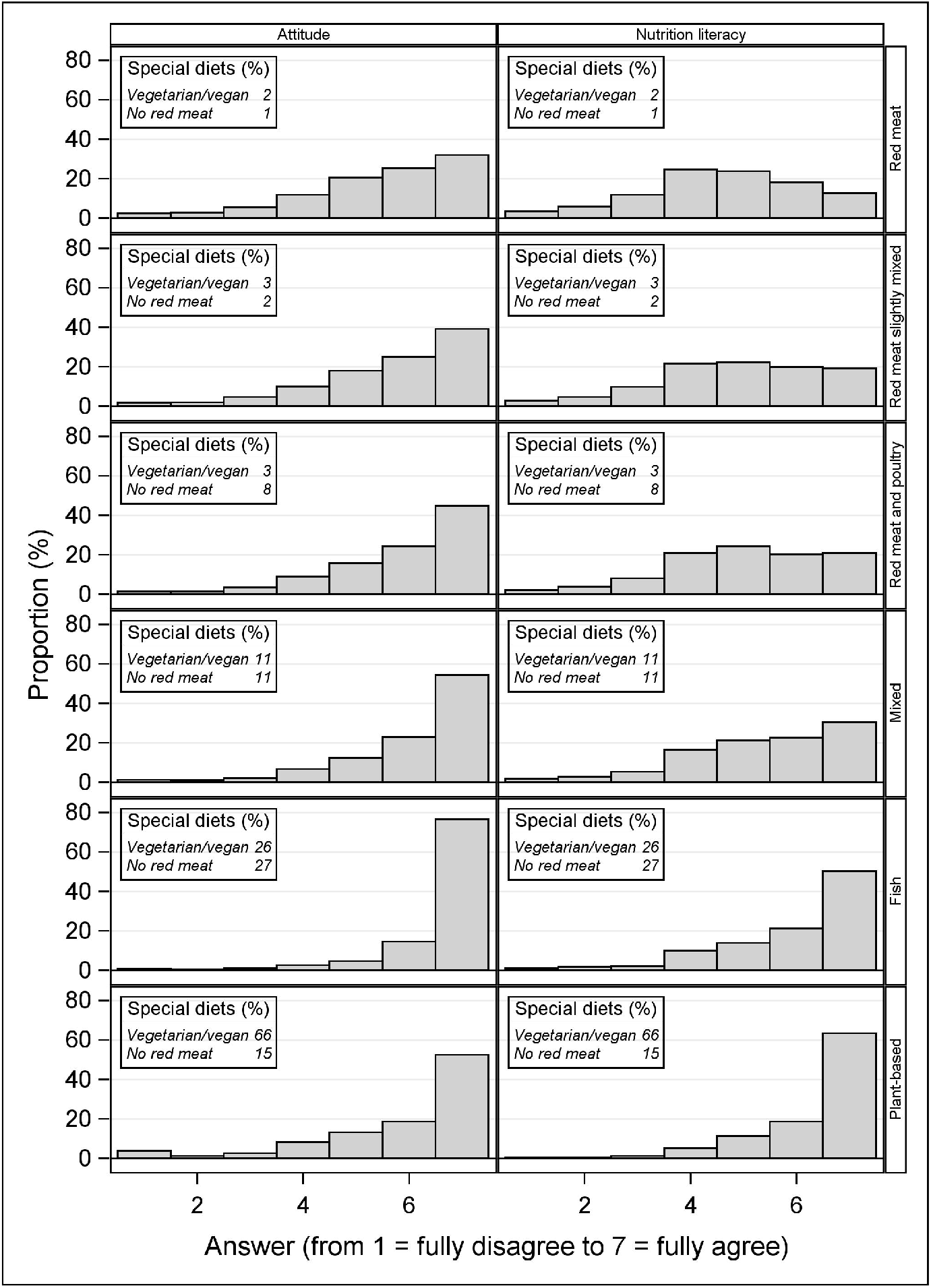
Frequency distribution (%) of attitude (“To promote health, the consumption of fish should be increased and that of red meat and processed meats limited”) and nutrition literacy (“Recommended dietary changes include increasing the consumption of pulses”) within clusters.

An ordinal logistic model (Figure 3) considering the profiles ordered by their sustainability indicated a pronounced trend across education levels; those with higher education tended to have more sustainable purchase profiles. An inverse yet much weaker relation was observed for household income; participants with the lowest household income tended to have more sustainable profiles. The analysis also confirmed that households with children tended to have less sustainable profiles than single or two-adult households. Attitude and nutrition literacy seemed to act as intermediate factors for gender; women had somewhat more sustainable profiles than men, and attitudes and nutrition literacy accounted for approximately half of this difference. The model suggested a generation gradient, even after full adjustment. The youngest age group stood out from the rest, showing a tendency towards more sustainable profiles.

**Figure 3.**
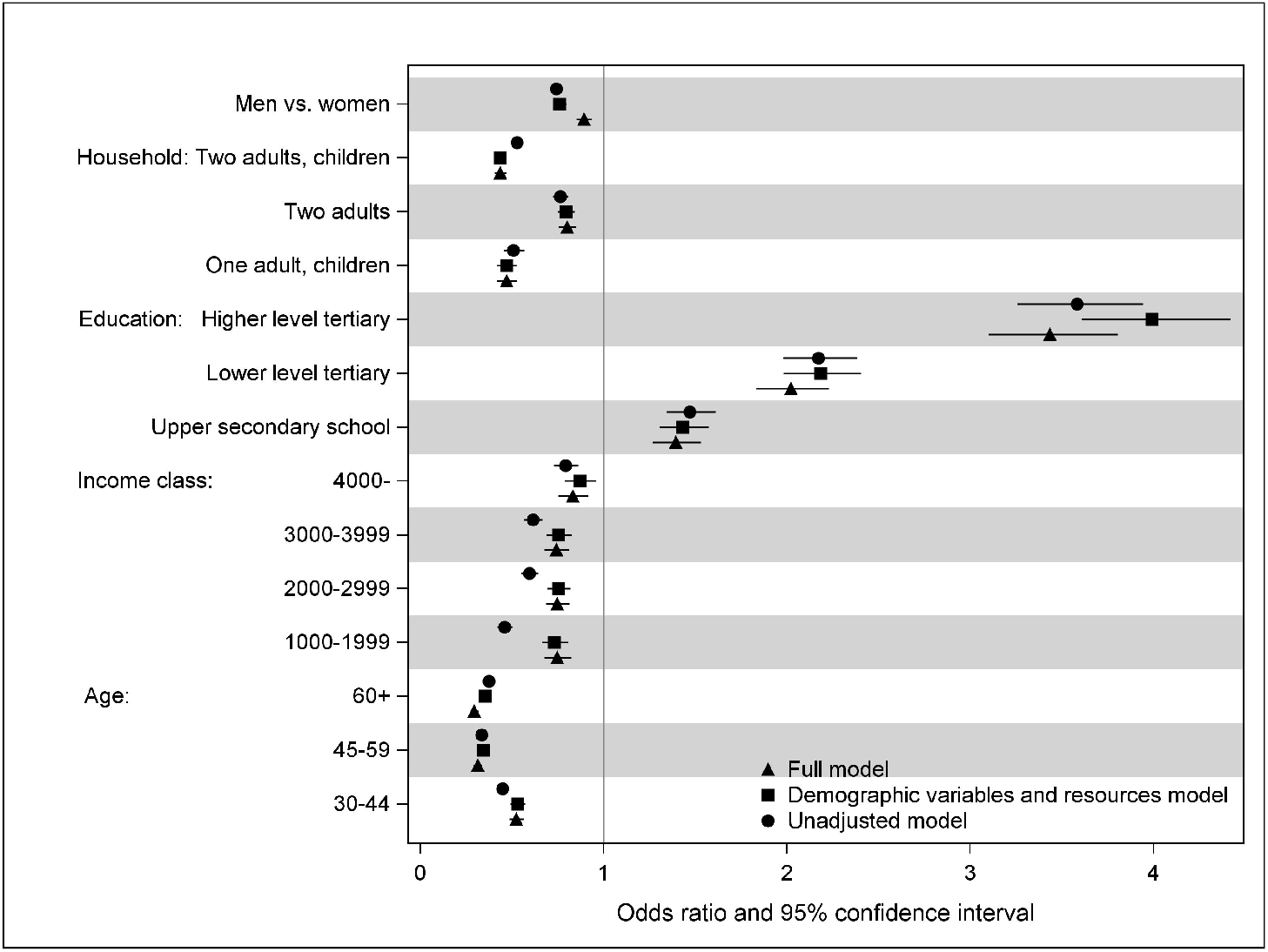
Odds ratios and their 95% confidence intervals from the ordinal logistic model. Odds ratios >1 indicate that the factor studied was associated with more sustainable, including a smaller proportion of red meat, purchase profiles. Reference class: single households and the lowest level in each factor. Full model includes all variables: demographic, resources, attitude, and nutrition literacy.

### Purchase prices

Table 3 presents the mean prices of the purchased protein sources. The average price paid was the lowest for plant-based foods, followed by poultry and red meat, and the highest for fish. There was considerable variation in purchase prices of products within all food groups. Among the ten most purchased fish and seafood products, the most purchased products were fresh rainbow trout and salmon, with prices ranging from 12.9 to 18.5 €/kg. The smallest prices were paid for fish sticks: 5.3 €/kg. Of red meat and processed meat, the most purchased product was minced meat, with a purchase price range of 5.0-8.2 €/kg, whereas cold meat cuts (11.3-12.7 €/kg) were bought with the highest prices. Traditional Finnish dishes of pea soup (2.6 €/kg) and spinach pancakes (3.9 €/kg) were the most purchased plant-based foods. The average purchase prices for the newly developed plant-based foods, such as pulled oats and vegetable sausage, ranged from 12.2 to 18.3 €/kg, but the purchase volumes of these products remained much smaller than those of traditional dishes. Participants with the red meat profile tended to pay lower prices on average for most of the protein sources than other participants. Those with the fish or mixed profile paid the highest prices.

**Table 3.** Mean (standard deviation) of participants’ purchase price (€/kg) by purchase profile and food group.

|  | **Food group** |  |  |  | **Overall** |
| --- | --- | --- | --- | --- | --- |
| **Profile** | Red meat and processed meat | Poultry and poultry dishes | Fish and seafood | Plant-based foods |  |
| Red meat | 7.7 (1.9) | 7.7 (2.0) | 13.5 (4.3) | 4.8 (2.1) | 8.0 (2.0) |
| Red meat slightly mixed | 8.4 (2.2) | 7.7 (2.2) | 14.2 (4.7) | 5.4 (2.3) | 8.7 (2.3) |
| Red meat and poultry | 8.9 (2.3) | 7.6 (2.1) | 14.2 (4.6) | 6.0 (2.4) | 8.7 (2.0) |
| Mixed | 9.3 (3.0) | 8.2 (2.6) | 14.9 (5.1) | 6.6 (2.6) | 9.7 (2.8) |
| Fish | 11.2 (6.5) | 9.1 (3.1) | 15.5 (5.0) | 7.5 (2.6) | 12.4 (3.3) |
| Plant-based | 9.6 (4.1) | 9.2 (3.5) | 14.7 (5.8) | 8.3 (2.1) | 9.2 (2.2) |
| **Total** | 8.5 (2.7) | 7.8 (2.3) | 14.2 (4.8) | 5.7 (2.5) | 8.8 (2.4) |

### Changes in purchase preferences

The estimated probabilities of preference persistence or transition from one week to another are displayed in Table 4. Despite positive attitudes towards changes from red meat to fish and increased consumption of pulses, the probability of persistence of red meat preference remained steady and by far the strongest (82%). This was followed by persistence of plant-based food preference (53%). The persistence of all preferences and the departure and entry probabilities for the two above-mentioned preferences over the follow-up period are displayed in Figure 4. The probabilities were fairly stable over time; however, a slight increasing trend in the persistence of plant-based food preference was observed. Departure from red meat was the most likely towards poultry, followed by fish and plant-based foods. The probability of entry to plant-based food preference was the reverse; the most common entry was from fish preference, followed by poultry and red meat preferences. Relative differences in the transition probabilities were two-or even threefold across food groups.

**Table 4.** Persistence (shaded cells) and transition probabilities from a purchase preference one week to a preference in the subsequent week. Missing preferences are not displayed; hence, the row probabilities do not add up to unity.

|  | To red meat | To poultry | To fish | To plant-based |
| --- | --- | --- | --- | --- |
| From red meat | 0.82 | 0.08 | 0.05 | 0.03 |
| From poultry | 0.35 | 0.48 | 0.08 | 0.05 |
| From fish | 0.31 | 0.11 | 0.46 | 0.08 |
| From plant-based | 0.24 | 0.08 | 0.10 | 0.53 |

**Figure 4.**
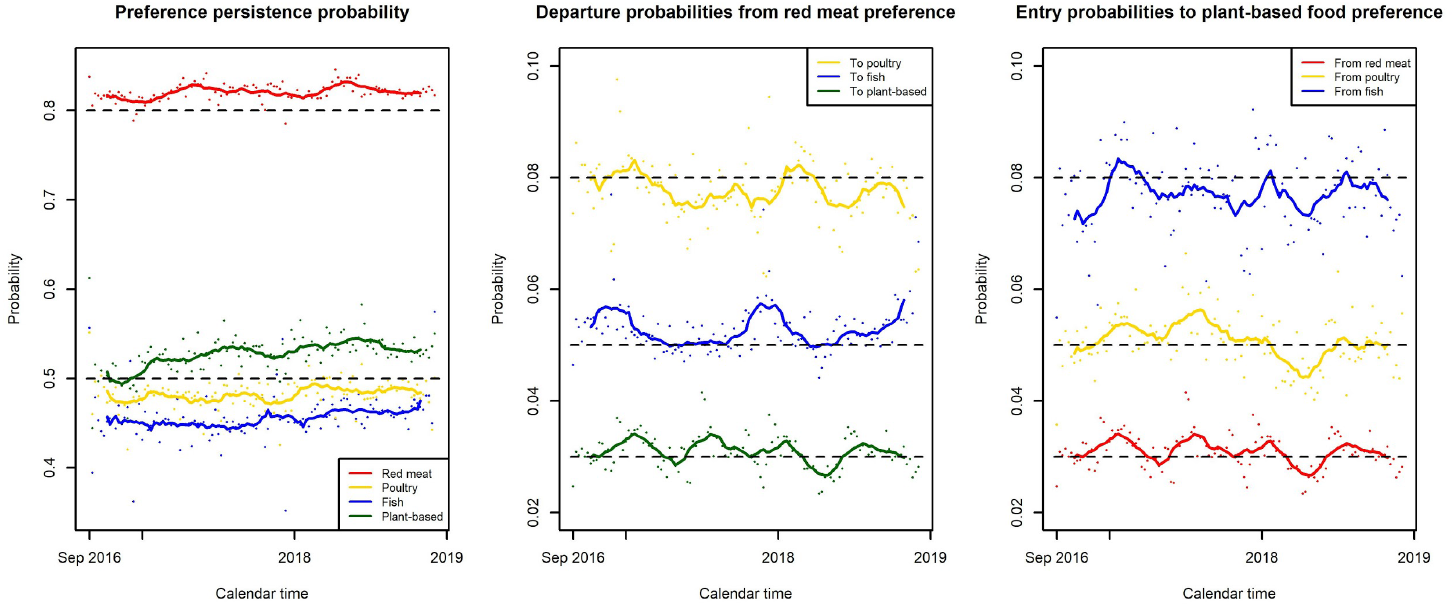
Estimated probabilities of persistence of, departure from, and entry to a preference from one week to the next by calendar time. Points indicate probabilities by week, and solid lines their 12-week moving averages.

## Discussion

Our analyses of large Finnish grocery purchase data identified six purchase profiles with different emphases on red meat, poultry, fish, and plant-based foods. Overall, red meat dominated the purchases. Considerable variation existed in purchase prices within all food groups, with the lowest average price paid for plant-based foods. Red meat and plant-based preferences showed the highest persistence. Transitions from red meat were most likely to shift towards other animal foods, favoring poultry over fish and fish over plant-based foods.

Our approach confirms recent findings from more traditional analyses (e.g. 31, 32); despite the media hype and accumulating evidence supporting sustainable protein sources, red meat preference and low regard for plant-based options are highlighted in purchase and consumption profiles. Regardless of the slow rise in the proportion of vegan, vegetarian, and red meat-free diets, the prevalence of different vegetarian diets remains modest in Finland (33) and in other Western countries (22, 34). Also contrary to the impression given by public debate, there was no evidence of polarization to opposite extremes (red meat or plant-based foods) on the preference spectrum; the majority (70%) of participants demonstrated somewhat mixed purchase profiles.

Substituting red meat with fish, but not with poultry, may be associated with cardiovascular health benefits (4), and there are sustainable fish choices available (35). Importantly, there seems to be room for improvement in each purchase profile. Even those with plant-based food profiles appeared to prefer red meat over fish. Future efforts need to be placed to accelerate substituting meat with sustainable and affordable fish choices.

Currently, conceptual indicators and metrics for the economic and social domains of the sustainable diet are the least developed (36). We particularly stressed these domains and observed that lower purchase prices tended to appear among less sustainable purchase profiles. Within the food groups, however, the average price paid was the lowest for plant-based foods and almost triple that price for fish and seafood. Thus, more expensive products are being purchased from the fish group or the variety of cheaper fish products is not as wide or appealing. Differentials in the price of healthy and unhealthy foods and diets may contribute to health inequalities (36). Based on our findings, price may act as a barrier to fish purchases, but did not prevent purchase of plant-based foods. Participants with the lowest household income had the most sustainable profiles, as was observed also in France (22). Shifting from the current meat and protein-oriented diets to the current national dietary guidelines has been projected to save more than one-third of the daily food expenditure primarily due to replacement of expensive meat products with cheaper plant-based foods (10, 37). Previously, we demonstrated socio-demographic disparities in food choice motives, with higher priority placed on familiarity and price in people with lower socio-economic position and among men (38). Vainio et al. (39) found lower price to be an important food choice motive among current beef consumers, in agreement with our observation that they tended to buy cheaper products.

It is noteworthy that the majority of participants agreed that the consumption of fish and pulses should be increased and that of red meat and processed meat decreased. However, attitudes and knowledge had not actualized in purchase behavior, as the proportions of fish and pulses in the purchase profiles were relatively small. This is in line with a well-known gap between stated attitudes or intentions and actual behavior (40). To increase sustainability of consumers’ food purchases, multiple actions that target both environmental (including price) and more individual level behavioral determinants are needed. Importantly, of interventions targeting conscious determinants of behavior, self-monitoring and individual lifestyle counselling interventions have been most promising in reducing actual consumption of meat (41).

We found that the most important resources in choosing more sustainable profiles were education and stage of family life. Similarly to previous studies (17, 18, 22, 23, 33), the red meat profile was common among less educated persons and men, but also among middle-aged participants with a family. The observed differences between the sexes could have been slightly diluted by the fact that purchases were made for an entire household. The youngest age group seemed to have replaced red meat to a large extent with poultry and plant-based food alternatives, but not with fish. This may indicate a slow intergenerational change.

Our results indicate a slow road with tiny steps from meat dominance to more sustainable diets. This was reflected by our findings of a mixture of prevalent profiles, with the highest persistence of the red meat preference. The most likely departure from red meat was towards poultry, followed by fish and plant-based foods. This supports earlier findings of the tendency to use a hierarchy of foods when substituting meat; food is often substituted on a plate by another familiar product with a similar component structure (42, 43). These hierarchical steps are more likely than leaps, especially among the older generation.

The step-by-step approach should be valued when designing health communication and steering methods. Switching to poultry and fish must be presented as an attractive, affordable and socially valued option since a direct switch to plant-based foods might be too much of a leap for most consumers. While information and education are critically important means, shifting attention also to consumers’ everyday practices and habits is important to generate long-lasting shifts (44).

Shifting towards sustainable food choices requires multilevel national and international collaboration between all stakeholders (45). Dietary guidelines that reflect the latest evidence on healthy eating complemented with broader and more explicit criteria of sustainability can be important macro-level tools for improving health and environmental sustainability (11, 45) Taxation may be an effective way to enhance major changes, given that alternative products are markedly cheaper (46).

Sustainability goals also call on food retailers to engage in social responsibility by extending their focus from addressing consumers’ preferences to the common good. Grocery store interventions that manipulate price and item availability, or suggest swaps, have an impact on purchase behavior (47). Food retailers’ use of loyalty card data to uncover consumer preferences could be utilized to design effective promotional activities, e.g. individual feedback, or tailored price discounts that facilitate consumers’ change towards more healthy or sustainable preferences. This could help the food retailer to build competitive advantage to other retailers (48). Indeed, several aspects may have already accelerated the transition towards a more sustainable diet since our data collection in 2016-2018. The market value for plant-based products has been estimated to grow 7% annually in 2019-2021 (49), and the supply and affordability of more sustainable foods have most likely improved. An encouraging finding was that we observed a slight increasing trend within the 2.3-year follow-up in the persistence of the plant-based food preference. Moreover, disruptions to food systems caused by the COVID-19 pandemic create opportunities to drive longer term transformation (50).

Strengths of this study include a remarkably large dataset and a long and detailed follow-up period. Automatically collected data such as these will likely become complementary to self-reported data in future evaluations of health and sustainability issues, and it is therefore important to develop visualization and analysis methodology to use these data efficiently. The choice of methodology from another field of science may provide new insight into unconventionally structured data on food consumption or such surrogates as loyalty card holder purchases (51).

We have demonstrated that loyalty card data on grocery purchases reasonably well reflects the self-reported food and drink consumption of the adult population (52, 53). However, purchase volume does not correspond to consumed volume. The preparation of food may lead to marked changes in weight (54). Consequently, the share of plant-based foods may be underestimated and that of meat and fish overestimated. We believe that the profiles identified in our study could be generalizable to similar consumer environments such as the Nordic countries, where consumer culture is influenced by the Nordic societal model of strong welfare system, relatively open markets, and relatively egalitarian stance to social differentiation (55).

We focused on four protein sources with clearly distinct environmental and health impacts. Increasing the consumption of more climate-neutral foods, such as whole grains, could also contribute to meeting environment-focused sustainability targets without compromising the healthiness of the whole diet (3). In future studies, the whole-diet approach should be preferred. We applied single questions to measure nutrition literacy and attitude towards a topic, which were insufficient for assessment on a broader level. Further research should apply well-defined and theoretically grounded measures (16). A limitation of the study is that the participants were more likely women and more highly educated than the general adult Finnish population (27). A sensitivity analysis using a full re-weighting (27) showed that our results on the prevalence of red meat and plant-based profiles could be slightly underestimated and overestimated, by five and two percentage points, respectively. Other profiles had a similar prevalence in the two analyses.

To conclude, although red and processed meat dominated the purchase preferences and showed the highest persistence, there are promising signs for a shift towards more sustainable diets. Consumers’ attitudes are set for a change, and many plant-based foods are affordable regardless of income. Reducing the price difference between the most commonly purchased red meat and fish products could further accelerate this development. It is important to ensure ways for today’s young “greener” generation to maintain and strengthen the commitment to plant-based foods as they enter middle-age and face a hectic family life. Societal incentives for sustainable food choices seem most crucial at transition stages of life course and for the less educated. The world’s future protein supply is an extremely challenging interplay between ecological, social, cultural, and economic sustainability and the necessary nutritional demand for high-quality protein. Up-to-date individualized grocery purchase data offer a means for developing, implementing, and monitoring progressive policies towards a healthy and sustainable food system.

## Data Availability

Program codes and their output files will be available from the corresponding author. Data can be analyzed in collaboration with the research team on a reasonable request.

## Acknowledgements

We thank the S Group for collaboration. We are also grateful to the loyalty card holders who provided consent for the use of their loyalty card data in this research project.

## Author contributions

ME, HS, MF and JN were involved in the data collection and transfer. ME and JN were responsible for the design of the study, wrote the first draft of the manuscript, and had primary responsibility for the final content. JN was responsible for the analyses of the study. All authors participated in commenting and critically revising the first draft of the manuscript, and read and approved the final version.

## Conflict of interest

HV has received a fee from the S Group. The collaboration included offering professional advice to influencers and writing a blog post with regard to interpretation of nutrition calculator in S Group’s mobile app. MF is a member of the S Group’s Advisory Board for Societal responsibility. The membership is without any compensation. The authors declare no other relationships or activities that could appear to have influenced the present work.

---

**Supplemental Figure 1.**
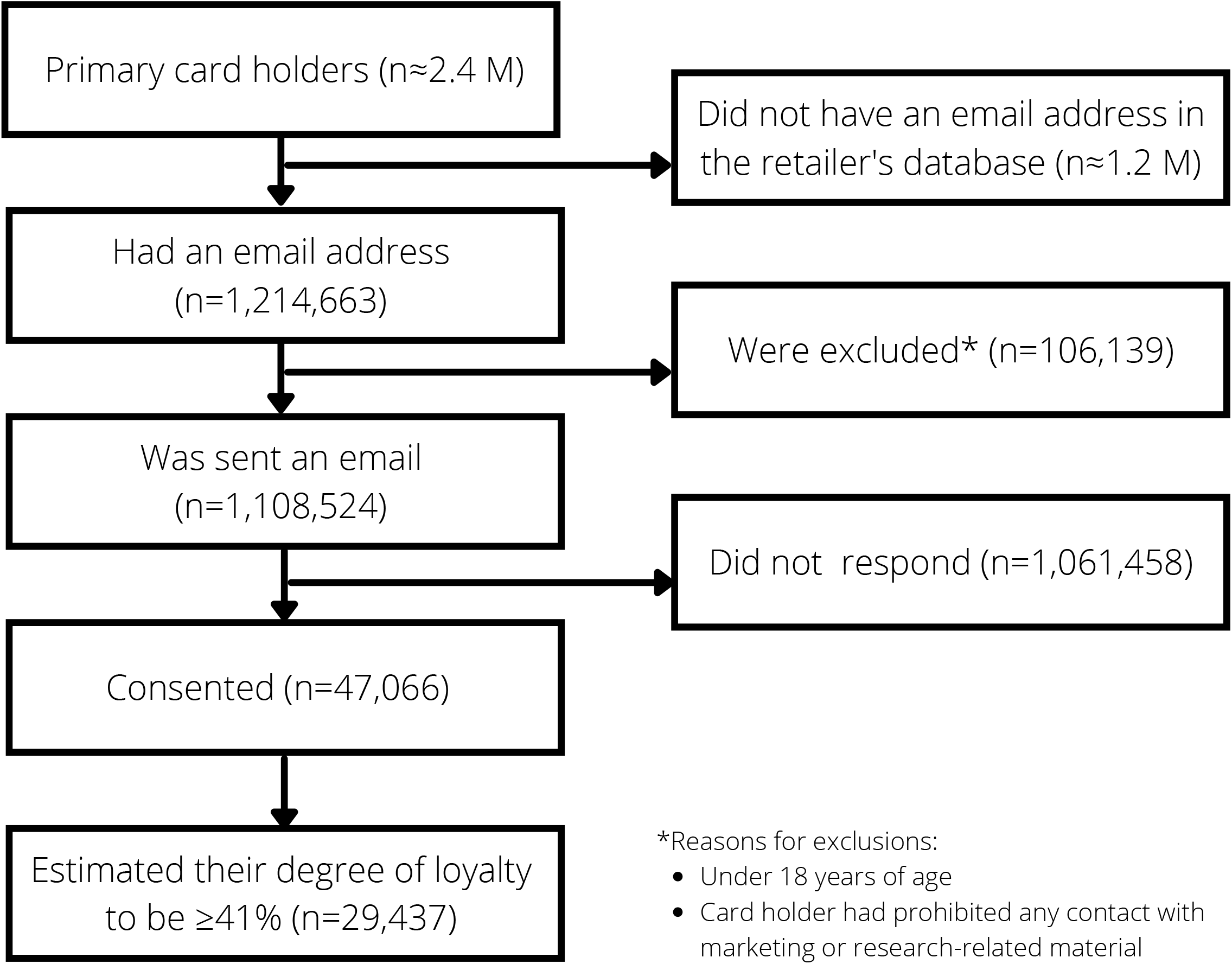
The flow of participants in the current study.

## Notes

**Sources of support:** The Academy of Finland (#309157, HK), the Emil Aaltonen Foundation (JM), the Yrjö Jahnsson Foundation (JM), and the Finnish Food Research Foundation (HV).

### Funding Statement

This study was funded by the Academy of Finland (#309157, HK), the Emil Aaltonen Foundation (JM), the Yrjo Jahnsson Foundation (JM), and the Finnish Food Research Foundation (HV).

### Author Declarations

The study was approved by the University of Helsinki Ethical Review Board in the humanities and social and behavioral sciences (Statement 21/2018). Each participant provided an informed consent electronically.

